# The Actively Secreted Plasma ExtraCellular Vesicle Troponin (ASPECT) Study: Circulating Troponin in Extracellular Vesicles Across Cardiovascular Disease Cohorts

**DOI:** 10.64898/2025.12.01.25340197

**Authors:** Michail Spanos, Priyanka Gokulnath, Aarush Singh, Christopher Azzam, Ronak Wakhlu, Claire Lin, Ana-Chrysa Maravelias, Timothy Oasan, Albree Tower-Rader, Megan Wasfy, Petr Jarolim, James L. Januzzi, Saumya Das

## Abstract

Cardiac troponin is essential for diagnosing myocardial infarction, yet high-sensitivity assays frequently detect troponin elevations in non-ischemic contexts, complicating clinical decision-making. We investigated extracellular vesicle-associated (EV) versus non-vesicular (NEV) troponin in plasma samples from 266 participants across acute and chronic heart failure, type 1 and type 2 MI, hypertrophic cardiomyopathy, end-stage kidney disease, healthy individuals, and exercise states. EV troponin was negligible in necrosis-dominant conditions (MI, kidney disease) but constituted up to 40–50% of total troponin in chronic heart failure or hypertrophic cardiomyopathy and nearly 100% in healthy or exercise cohorts. Unlike plasma troponin, EV troponin weakly correlated with natriuretic peptides or renal indices, suggesting a distinct release mechanism linked to chronic stress or physiological turnover. These findings highlight the potential for EV troponin to distinguish active, non-necrotic processes from acute injury. Further study may clarify its prognostic utility and refine current diagnostics and risk stratification.

**GRAPHICAL ABSTRACT:** 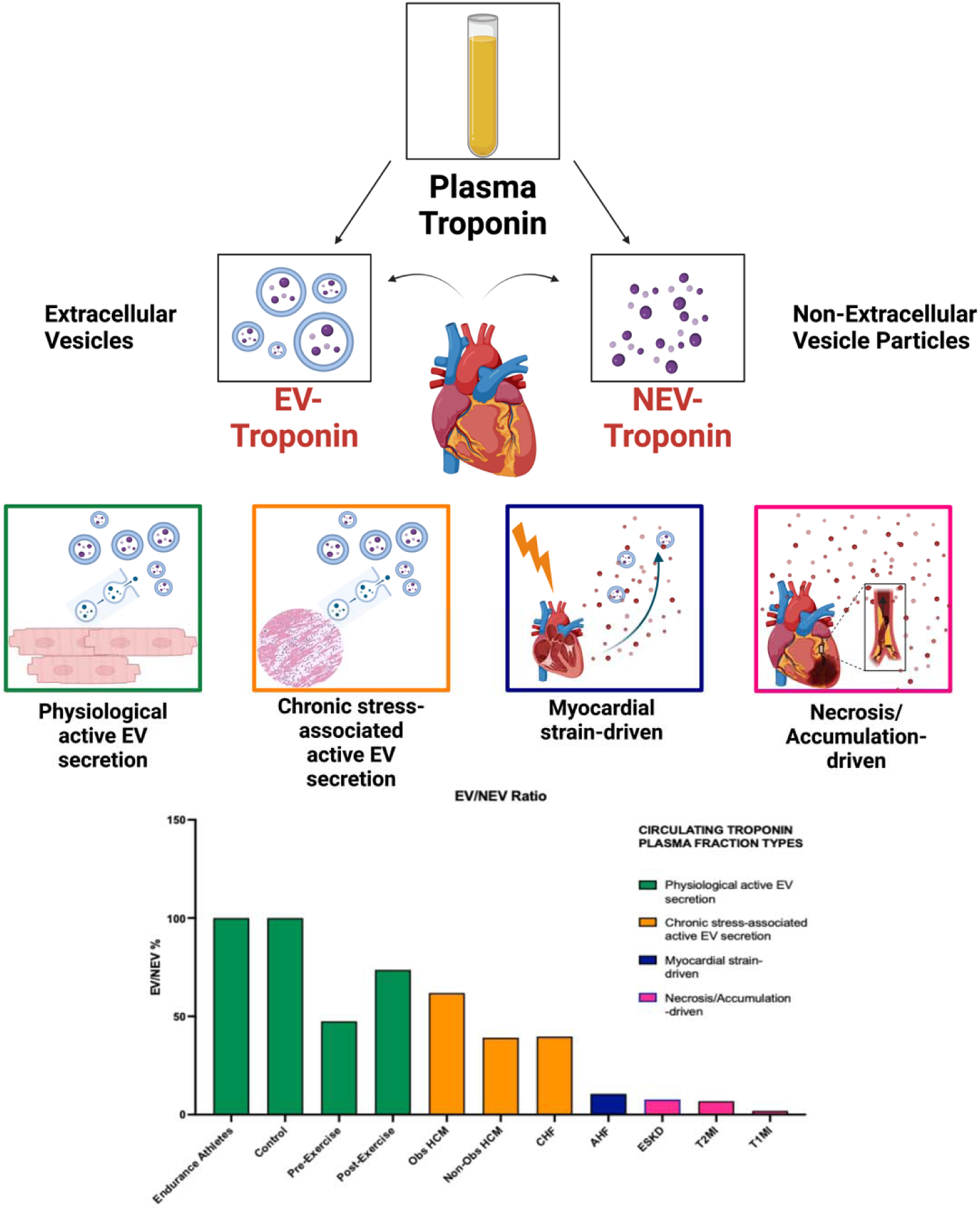

## INTRODUCTION

Cardiac troponins are important biomarkers for diagnosing myocardial injury, especially in the context of acute coronary syndromes (ACS). Acute myocardial infarction (MI) is defined by the presence of cardiac ischemia in the presence of a rise and/or fall of troponin (Thygesen et al., 2018). These proteins, key components of cardiomyocyte contractile apparatus, are localized mainly in the sarcomere but a small percentage is found in the cardiomyocyte cytoplasm. Sarcomeric and cytosolic troponin are released into the bloodstream when myocardial injury causes compromise of cell membrane integrity. In this context, increased concentrations of troponin may be detected in the circulation within minutes of cardiomyocyte injury particularly when high-sensitivity cardiac troponin (hs-cTn) assays are used. These assays have revolutionized the management of ACS, enabling more rapid MI diagnosis and enhanced risk stratification (Mohammed & Januzzi, 2010).

Although hs-cTn represents the global standard for diagnostic evaluation of acute MI, with use of hs-cTn assays, elevated troponin concentrations have been recognized in a significant number of individuals who do not have acute MI. Such non-MI related troponin excursions are frequently observed in a variety of conditions that involve myocardial stress, damage, or systemic insult without classic ischemic injury (Akbas, 2024; Eggers et al., 2019; Januzzi et al., 2010; Park et al., 2017). Such states associated with abnormal troponin without MI include chronic kidney disease (CKD), heart failure (HF), certain cardiomyopathies, or even extreme exercise. This observation suggests that troponin release from the cardiomyocyte might be attributed to mechanisms other than myocardial necrosis (Eggers et al., 2019; Park et al., 2017). Besides leading to diagnostic ambiguities (Akbas, 2024; Ilva et al., 2010) the presence of “non MI” related troponin changes imply the potential for a different mode of release of the biomarker into circulation. Understanding how troponin might leave the cardiomyocyte besides due to cardiomyocyte necrosis might inform means by which to understand such abnormal circulating concentrations in those without MI.

Small extracellular vesicles (sEVs, also known as exosomes and referred to as EVs in this manuscript) are small membrane-bound vesicles released by cells in the context of numerous cellular processes including inter-cellular communication. With their origin from the cytoplasm, EVs contain proteins, nucleic acids, lipids, and other metabolites. Given that a small percentage of troponin in myocardium is found in the cytosol, we previously examined the possibility that EVs might contain measurable concentrations of troponin. In a pilot study (Lennon et al., 2022) using advanced single-EV nanoscopy, troponin molecules were directly visualized within individual EVs for the first time, confirming that vesicular export of troponin is a biologically plausible pathway. This early work established proof-of-concept that troponin can be actively packaged and secreted within vesicles; however, it did not address the broader biological landscape, including the relative burden of EV release of this biomarker in the setting of stress, inflammation, and other pathophysiological stimuli as opposed to free troponin that is released from damaged or dying cardiomyocytes.

To date, no study has systematically examined the compartment-specific release of troponin, EV-encapsulated versus free-form troponin, across diverse cardiovascular conditions. Without understanding this compartment-specific biology, the clinical interpretation of troponin elevations in non-MI states remains limited. Here, we build upon our foundational observations by quantifying EV-associated and non–EV troponin across a spectrum of disease states, including acute MI (Type 1 MI, Type 2 MI), acute and chronic heart failure, obstructive and non-obstructive hypertrophic cardiomyopathy, end-stage kidney disease, physiologic exercise responses, and health. By establishing the degree and pattern of troponin compartmentalization, our study aims to clarify whether different mechanisms of troponin release underlie the diverse clinical scenarios in which elevated troponin is observed.

## MATERIALS AND METHODS

### Ethics and Study Population

The Mass General Brigham Institutional Review Board approved of all study procedures and all patients provided informed consent prior to blood draws. Plasma samples from 266 participants across ten cohorts were analyzed: chronic heart failure (CHF; n=25), acute HF (AHF; n=25), end-stage kidney disease (ESKD; n=25), Type 1 MI (T1MI; n=25), Type 2 MI (T2MI; n=25), obstructive hypertrophic cardiomyopathy (oHCM; n=29), non-obstructive hypertrophic cardiomyopathy (nHCM; n=31), stress-test cohort with paired pre- and immediate post-exercise draws (n=31 individuals), endurance athletes (n=25), and healthy controls (n=25). Type 2 MI was defined according to the Fourth Universal Definition of MI, based on supply-demand mismatch without evidence of acute plaque rupture. For T1MI and T2MI, blood was obtained at clinically indicated time points following diagnosis; timing was not standardized to symptom onset or peak troponin. Pre- and post-exercise samples were drawn immediately before and immediately after the clinical stress test; no delayed timepoint capture was performed. Moreover, pre- and post-exercise cohorts represent paired samples from patients referred for clinical stress testing. Specifically, the stress-test cohort contributed 62 samples (31 pre, 31 post) but 31 unique individuals. Some had subclinical/baseline ischemia or chronic coronary disease, but none had acute coronary syndrome at the time of sampling. Plasma was collected under a standardized protocol, and EVs were isolated prior to measuring troponin concentrations in both EV and NEV fractions.

### EV-Enriched Troponin Isolation and NEV Separation

Plasma was processed using size-exclusion chromatography (SEC) optimized for EV enrichment. Fractions enriched for EVs (fractions 1–5) were pooled as the EV fraction, while later fractions (6–20) were pooled as the non-EV (NEV) fraction. Both were concentrated by centrifugal filtration to comparable volumes. EV identity was confirmed both quantitatively by nanoparticle analysis (biometric characteristics and size) and qualitatively by Western blot for canonical markers (CD81, ALIX). This approach ensured clear separation of vesicular versus non-vesicular troponin. Technical optimization details are provided in the **Supplementary Method: Developing and Optimizing the Troponin-Rich EV Isolation Method**.

### Western blot

The samples were lysed via sonication, and protein concentration was measured using a BCA protein assay kit (Thermo Fisher, Waltham, MA, USA), loaded on SDS-PAGE gel, transferred to PVDF membrane, which was blocked with 5% BSA in TBST, and incubated with primary antibody overnight and HRP conjugated secondary antibody for 1 hour. The blots were developed (**Supplementary Figure 2**) using SuperSignal® West Femto Maximum Sensitivity Chemiluminescent Substrate (Thermo Fisher, Waltham, MA, USA).

### Troponin detection

Cardiac troponin T concentrations were measured on the Roche cobas (Cobas e601) analyzer using the 5th generation high-sensitivity immunoassay with electrochemiluminescent detection (Roche Diagnostics, Indianapolis, IN, limit of quantitation: 6 ng/L). This FDA-approved platform is the standard assay used in routine inpatient care across most U.S. hospitals, ensuring direct clinical relevance of measured concentrations. For values below detection, concentrations were modeled at 50% of the limit (3 ng/L), primarily affecting healthy control and athlete samples.

### Statistical Tests

Both ANOVA and Kruskal-Wallis tests were used to evaluate overall differences in EV/Plasma ratios across disease groups. Paired t-tests and post-hoc Kruskal-Wallis comparisons were conducted to identify significant pairwise differences between groups. The EV/Plasma and EV/NEV ratios were calculated to assess the relative distribution of troponin in EVs versus non-EVs, with values log-transformed to approximate normality for parametric testing. Pearson correlation coefficients were computed to examine associations between EV troponin and clinical markers, including NT-proBNP, creatinine, and eGFR. All statistical analyses were performed using GraphPad Prism, Python, and R. To maintain data integrity and statistical validity, troponin values below the limit of detection (LOD) were modeled as 50% of the LOD (3 ng/L, since the LOD is 6 ng/L). Values near or below the limit of detection were primarily observed in healthy controls, endurance athletes, and pre-/post-exercise samples, but were rare in acute disease groups (MI, AHF, ESKD). This did not impact the interpretation of high-concentration states. Since the HCM cohort had limited sample availability, only EV and NEV troponin were measured, and the EV/Plasma ratio could not be calculated. Instead, the EV/NEV ratio was determined to provide a relative assessment of vesicular vs. non-vesicular troponin distribution within this group.

## RESULTS

### 1. Cohorts’ description

As noted, to determine the relationship between EV-associated troponin and how it contributes to the total measured troponin levels, we quantified EV, NEV, and total hs-cTn concentration in the plasma of different cardiovascular, renal diseases and physiologic states including Type I and Type II MI, AHF, CHF, ESKD, oHCM, nHCM, Pre- and Post-exercise and Endurance athletes as well as controls. Pre- and post-exercise cohorts reflect paired samples from patients referred for stress testing, some of whom had baseline/subclinical CAD but no ACS.

The cohort included predominantly male participants, with a mean age ranging from 61 to 72 years across groups, with the youngest group being T1MI (median 61 [IQR: 53–66] years) and the oldest group being T2MI (median 72 [IQR: 59–79] years) (**Table 1**). Left ventricular ejection fraction (LVEF) was notably reduced in patients with AHF and CHF, with median values of 35% [IQR: 27–50] and 35% [IQR: 35–61], respectively, with the rest of the study participants having an LVEF in the normal range. As expected, NT-proBNP levels were markedly elevated in individuals with AHF (median 6329 [IQR: 3192–11461] pg/mL) and ESKD (median 18308 [IQR: 5598–55008] pg/mL), reflecting significant cardiac stress. Participants in the T1MI and T2MI groups had lower NT-proBNP levels (328 [IQR: 110–972] pg/mL and 629 [IQR: 388–1422] pg/mL, respectively), while HCM groups exhibited moderate elevations (918 [IQR: 396–1704] pg/mL in nHCM and 408 [IQR: 168–2271] pg/mL in oHCM). Creatinine levels also followed the expected pattern, with ESKD patients showing the highest values (median 5.62 [IQR: 4.52–7.89] mg/dL), while other groups had values within normal or near-normal ranges.

**Table 1:**
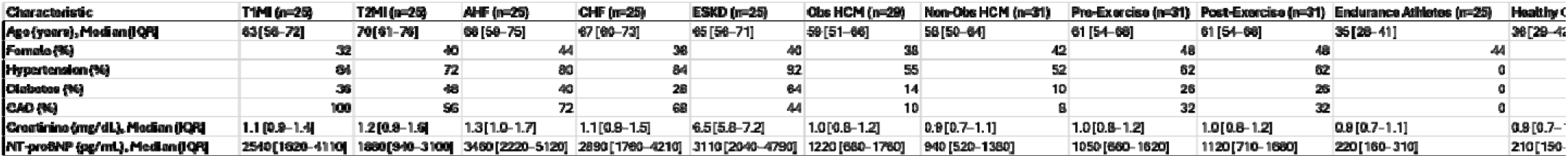
Baseline characteristics of the cohort. Baseline characteristics of the cohort, including demographic information, clinical variables, and relevant biomarkers for each group. Pre- and post-exercise cohorts reflect paired samples from patients referred for stress testing, some of whom had baseline CAD but no ACS. HCM samples (obstructive and non-obstructive) had lower available plasma volumes, which limited some downstream assays

Overall, hs-cTnT concentrations were, as expected, highest in T1MI followed by T2MI, AHF, and ESKD (**Supplementary Figure 3C)**. Specifically, hs-cTnT concentrations were significantly elevated in T1MI (median 871 [IQR: 563–1627] ng/L). By contrast, participants in the pre /post exercise Stress Lab cohort and the endurance athlete group had several troponin values below the limit of detection (LOD <6.0 ng/L).

### 2. Comparison of EV/Plasma and EV/NEV Ratios Across Disease Groups

We first measured the levels of troponin as measured by the hs-cTnT assay in the total (unprocessed) plasma samples, the EV fraction, and the non-EV fraction. Then, to identify which disease states were associated with higher proportions of EV-associated troponin, we calculated the EV/NEV and EV/Plasma ratios (**Figure 1A** and **1B**, respectively). A summary of median values and interquartile ranges is provided in **Table 2**, which shows disease-specific differences in troponin distribution across plasma, EV, and NEV compartments. These differences are further visualized in **Figure 3A and 3B**, emphasizing the continuum of secretion patterns across physiological and pathological states.

**Figure 1.**
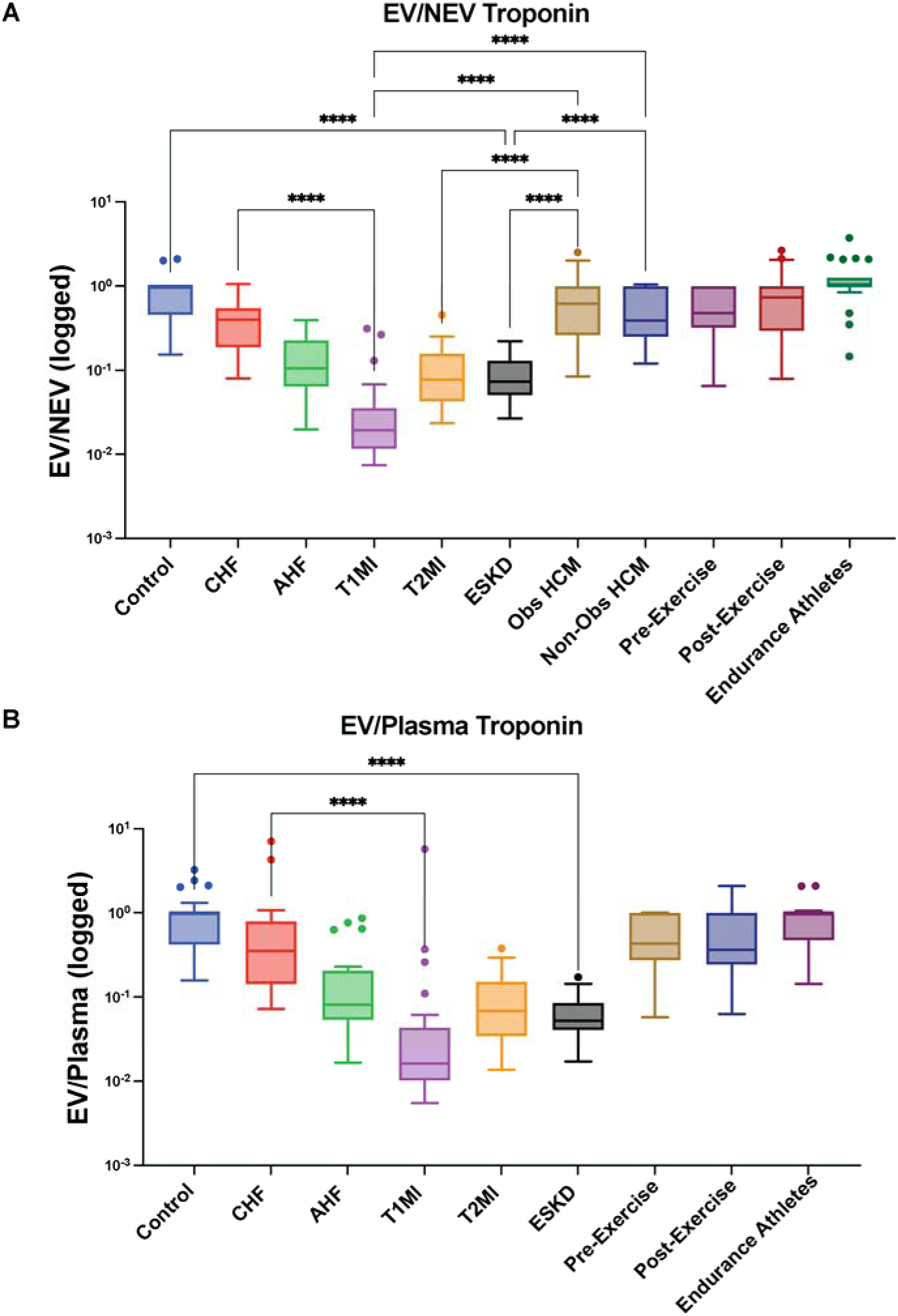
**A**. Extracellular Vesicle (EV) / Non- Extracellular Vesicle (NEV) Troponin ratio **B**. EV/ Plasma Troponin ratio across different physiological and pathological groups, including Endurance Athletes, Healthy Controls, Pre- and Post-Exercise, Hypertrophic Cardiomyopathy (HCM), Chronic Heart Failure (CHF), Acute Heart Failure (AHF), Type 1 Myocardial Infarction (T1MI), Type 2 Myocardial Infarction (T2MI), and End-Stage Renal Disease (ESRD). EV indicated by Extracellular Vesicle Fraction, and NEV indicated by Non-Extracellular Vesicle Fraction of 1mL Plasma isolated using size exclusion chromatography. The most significant comparisons are shown with * p<0.0332 ** p<0.0021 ***p<0.0002****p<0.00001where p is calculated using Kruskal-Wallis test using GraphPad Prism. For the entire pairwise comparisons using Dunn’s test (after adjustment) please refer **Supplementary** Figure 4 for EV/NEV (**A**) and EV/Plasma (**B**) comparison between the different groups.

**Figure 2.**
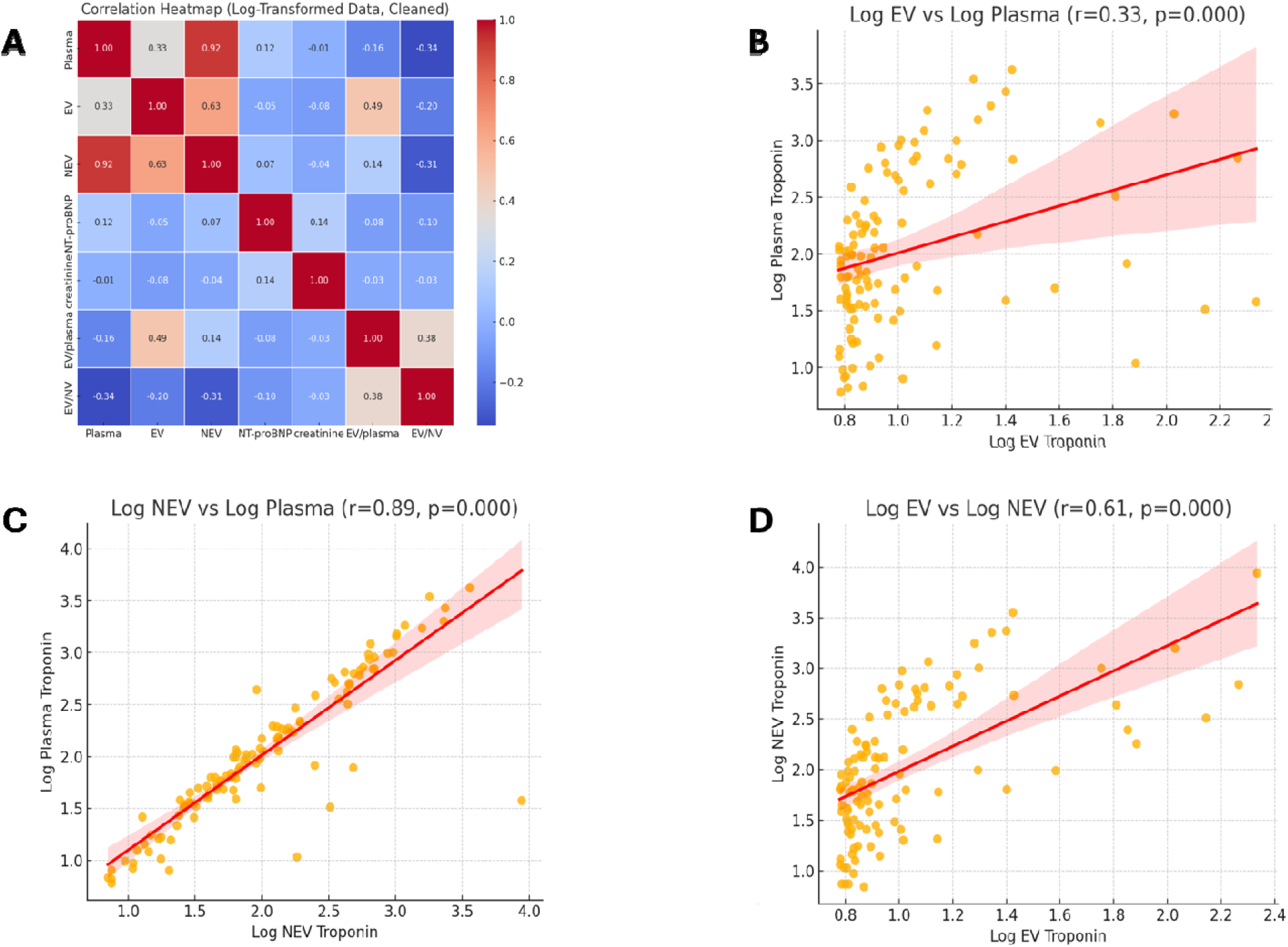
**A.** Correlation matrix showing the Spearman’s correlation coefficient between Plasma Troponin (Plasma), Extracellular Vesicle Troponin (EV), Non- Extracellular Vesicle Troponin (NEV), NT-proBNP, Creatinine and Ejection Fraction (EF) from the entire cohort. **B.** Correlation between EV and Plasma Troponin. **C.** Correlation between NEV and Plasma Troponin **D.** Correlation between NEV and EV Troponin from the cohort. All values are calculated using Pearson’s correlation coefficient using GraphPad Prism with ****p<0.0001.

**Figure 3.**
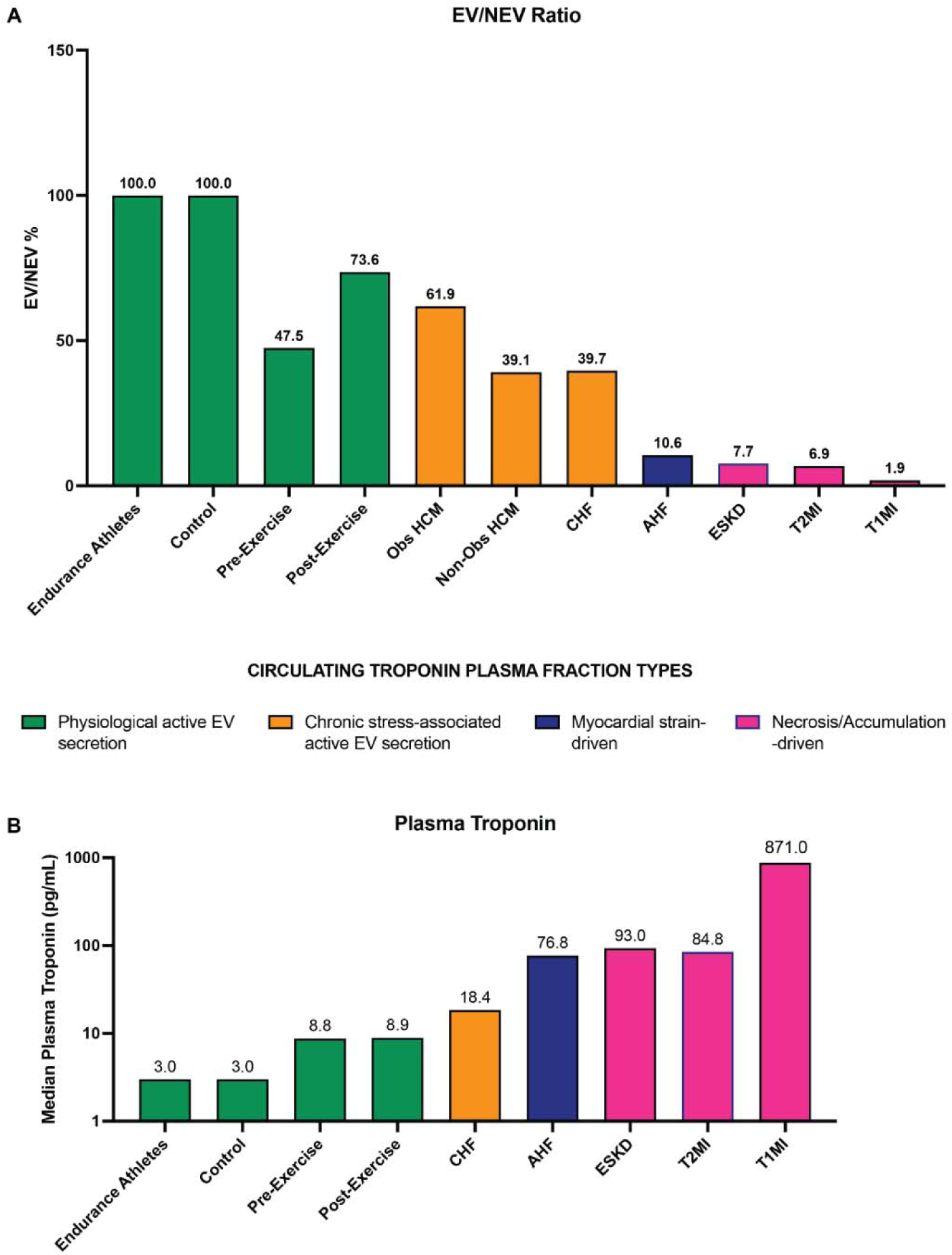
Circulating Troponin Secretion Modes Based on EV / NEV Troponin Ratio & Plasma Troponin Levels. **A.** EV (Extracellular Vesicle) / NEV (Non-Extracellular Vesicle) Troponin ratio represented as percentage across different physiological and pathological groups, including Endurance Athletes, Healthy Controls, Pre- and Post-Exercise, Hypertrophic Cardiomyopathy (HCM), Chronic Heart Failure (CHF), Acute Heart Failure (AHF), Type 1 Myocardial Infarction (T1MI), Type 2 Myocardial Infarction (T2MI), and End-Stage Renal Disease (ESRD). High EV/NEV ratios in athletes and controls indicate physiological secretion, whereas chronic stress-associated conditions (CHF, HCM) show moderate EV-associated troponin release. Myocardial strain (AHF) exhibits mixed secretion, while necrosis-driven conditions (T1MI, ESRD) show minimal EV involvement. **B**. Plasma troponin levels (pg/mL) in the same groups highlight differences in release mechanisms. Physiological groups exhibit low plasma troponin, while chronic stress (CHF, HCM) shows moderate elevations. Myocardial strain (AHF) results in higher levels, and necrosis-driven conditions (T1MI, ESRD) show the highest plasma troponin concentrations. Color coding: **Green** – Physiological EV secretion, **Orange** – Chronic stress-associated EV secretion, **Blue** – Myocardial strain-driven, **Pink** – Necrosis/Accumulation-driven. EV and NEV fractions were isolated using size exclusion chromatography.

**Table 2:**
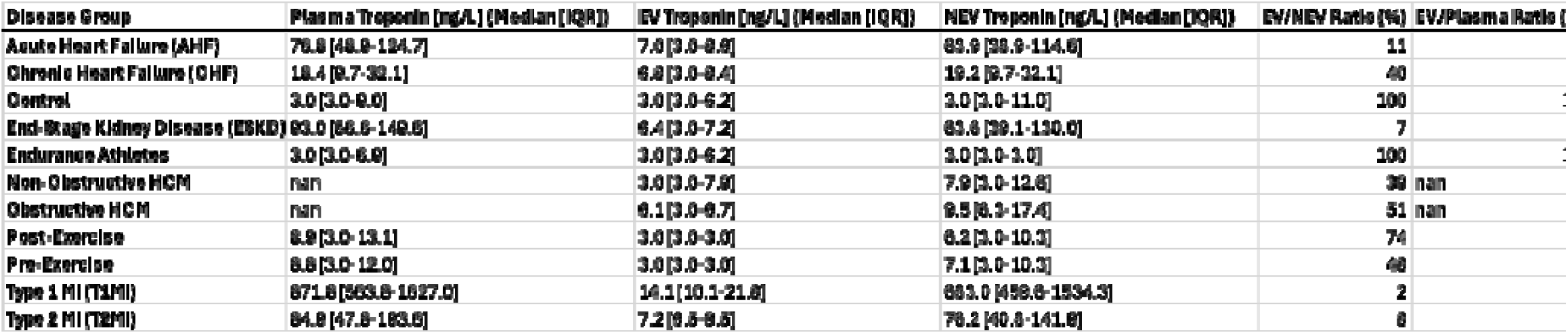
EV and NEV Troponin measurements of different groups in this cohort. EV and NEV troponin measurements across different groups in the cohort, showing the median and interquartile range (IQR) of plasma, extracellular vesicle (EV), and non-extracellular vesicle (NEV) troponin levels. The table also includes EV/NEV and EV/Plasma ratios to illustrate the relative distribution of troponin in each compartment.

In those with T1MI, plasma troponin concentrations were markedly elevated, with an EV/Plasma ratio of only 2% and an EV/NEV ratio of 2% as well, indicating that the vast majority of troponin was present in the free (non-EV) fraction. Similarly, those with T2MI demonstrated high plasma troponin levels (median 84.9 [IQR: 47.8–193.6] ng/L; **Table 2**), with an EV/Plasma ratio of 8.5% and an EV/NEV ratio of 9.5%, slightly higher than T1MI indicating that while in both T1MI and T2MI, troponin is released predominantly via passive leakage from necrotic cardiomyocytes, T2MI exhibits higher active vesicular secretion.

Individuals with ESKD demonstrated the next highest plasma troponin levels, (median 93.0 [56.6–149.6] ng/L; **Table 2**). Curiously, despite the high plasma levels, ESKD had a low proportion of EV-associated troponin (EV/Plasma ratio = 5%, EV/NEV = 7%), suggesting that troponin accumulation in these patients is driven more by cell death or reduced renal elimination rather than an active vesicular secretion process.

Among those with AHF (median plasma troponin 76.8 [48.9–124.7] ng/L; **Table 2**), the EV/Plasma and EV/NEV ratios were 8% and 11%, respectively, indicating that the majority of troponin released in this population was also in the non-EV fraction. In contrast, CHF and HCM demonstrated lower overall plasma troponin but higher proportions of EV-associated troponin. For example, CHF had a median plasma troponin of 18.4 [9.7–32.1] ng/L yet displayed an EV/Plasma ratio of 35% and EV/NEV ratio of 40% (**Figure 1A–B**), suggesting that vesicular secretion is a defining feature of chronic heart failure rather than necrosis. Similarly, HCM exhibited EV troponin levels of 3.0 [IQR: 3.0–7.9] ng/L in nHCM and 6.1 [3.0–10.1] ng/L in oHCM, with EV/NEV ratios of 39% and 51%, respectively, indicating that troponin levels in cardiomyopathies rely heavily on EV trafficking.

At the lowest end of the spectrum, healthy controls and endurance athletes had minimal circulating hs-cTnT, with median plasma levels of 3 ng/L (**Table 2**, **Figure 3B**). In these physiological states, nearly all circulating troponin was vesicle-associated, with EV/NEV ratios of ∼100% (**Figure 3A**). By contrast, the pre-exercise cohort demonstrated slightly higher plasma troponin (median ∼8.8 ng/L) but a lower EV contribution (EV/NEV ratio ∼47.5%). Following exercise, plasma troponin remained similar (median ∼8.9 ng/L), yet the EV fraction increased (EV/NEV ratio ∼73.6%), suggesting that exercise not only modestly elevates troponin but also dynamically shifts its mode of release toward vesicular secretion. Notably, the lower EV/NEV ratio in the pre-exercise group compared with healthy controls is likely explained by population differences: unlike healthy volunteers, these were patients clinically referred for stress testing, some of whom had baseline subclinical ischemia or chronic coronary disease. Such subclinical pathology may favor NEV release, reflecting ongoing myocardial stress even before exercise.

We next conducted a correlation analysis to further explore the relationship between EV and NEV troponin levels (**Figure 2A-D**) and other clinical markers, including NT-proBNP and kidney function markers (creatinine, eGFR) (**Supplementary Table**). Our results revealed that EV troponin levels were not significantly correlated with NT-proBNP (r = −0.046, p > 0.05), creatinine (r = −0.082, p > 0.05) or eGFR (r = −0.03, p > 0.05). In contrast, plasma troponin levels showed a strong correlation with NEV troponin levels (r = 0.92, p <0.001), while EV-associated troponin demonstrated a weak but statistically significant correlation with plasma troponin (r = 0.33, p = 0.027). These findings support the hypothesis that EV-associated troponin reflects a distinct biomarker pathway, separate from traditional mechanisms of troponin release.

In an effort to visualize differences between individuals in the various disease groups, the troponin results categorized by cardiovascular conditions or normal status were placed into four major groups (**Figure 3A–B**):

1. **Necrosis-driven troponin release or impaired clearance**, seen in MI and ESKD (where troponin is predominantly found in the free fraction due to cardiomyocyte necrosis or impaired clearance), with high plasma levels and low EV/NEV ratios
2. **Acute myocardial strain-associated release**, observed in AHF, with moderate EV/NEV ratios (∼11%), suggesting a combination of passive leakage and vesicular trafficking
3. **Chronic stress-associated vesicular secretion**, seen in CHF and HCM, which have high proportions of EV-associated troponin (EV/NEV = 40% in CHF, 39-51% in HCM), indicating that vesicular secretion may be a key feature of chronic cardiac stress rather than acute damage and
4. **Physiological vesicular secretion**, seen in healthy controls and endurance athletes, where plasma troponin is minimal, mostly being EV-associated, suggesting a role in normal myocardial turnover rather than a response to pathological stress.

## DISCUSSION

In this study, we compared EV–associated troponin with NEV troponin across a spectrum of disease states associated with elevated plasma troponin concentrations. We also examined EV and NEV troponin in healthy individuals. By quantifying the proportion of troponin contained within EVs relative to total plasma levels (EV/Plasma ratio) and to the NEV fraction (EV/NEV ratio), we aimed to discern whether different explanations exist for elevated hs-cTnT concentrations in these various states. Our findings confirm that depending on the disease state, a substantial percentage of circulating troponin may be contained within EVs. For example, heart muscle disease such as CHF and HCM as well as certain physiologic states (e.g., post-exercise) may be characterized by a higher proportion of EV troponin, whereas acute myocardial injury (T1MI and T2MI) and ESKD is dominated by NEV troponin. These results are the most conclusive to date that EV-based release of troponin may be meaningful and to our knowledge, these are the first results demonstrating heterogeneity in EV and NEV troponin in various disease states. Although troponin is an important diagnostic aid for the evaluation of individuals with suspected ACS, a vexing issue with this biomarker is the frequent observation of elevated concentrations in the absence of obvious acute MI. Abnormal troponin concentrations may create diagnostic uncertainty, leading to mis-diagnosis of MI (McCarthy et al., 2024). In the absence of acute MI, when abnormal troponin concentrations are frequently labelled “injury” but a plausible hypothesis is that other mechanisms for troponin release might be operative. Indeed, change in troponin concentrations are commonly seen in HF without coronary artery disease or in inflammatory disorders (Januzzi et al., 2012; Jeremias & Gibson, 2005; Kakihana et al., 2016; Tanindi & Cemri, 2011).

Our data highlight distinct EV/Plasma and EV/NEV troponin ratios across the clinical cohorts. For instance, individuals with CHF exhibited an EV/Plasma ratio of 35% and an EV/NEV ratio of 40%, indicating that a substantial proportion of the troponin in these patients was associated with EVs. Similarly, in HCM, a cardiomyopathy with genetic causes, the EV/NEV ratios were 51% in obstructive HCM and 39% in non-obstructive HCM, suggesting that troponin release in HCM involves both chronic myocardial strain and active vesicular trafficking. This pattern may represent persistent, active secretion tied to chronic stress, low-grade inflammation, or ongoing remodeling processes. These EV troponin elevations notably did not correlate with NT-proBNP, suggesting that EV troponin captures a facet of disease activity beyond mere wall tension or volume overload. In contrast, individuals with AHF had an EV/Plasma ratio of 8% and an EV/NEV ratio of 11%, values that are higher than in acute MI cohorts but significantly below those observed in CHF, implying that while acute decompensation may involve both active and passive release mechanisms, the latter predominate.

We observed that in T1MI, as expected the EV/Plasma ratio was only 2%, and the EV/NEV ratio was 2%, reinforcing the concept that troponin release in T1MI is almost exclusively non-vesicular, driven by extensive cardiomyocyte necrosis. This reflects the acute and catastrophic nature of coronary occlusion, where cell rupture leads to a massive spill of free troponin into circulation. Subtle differences between T1MI and so-called “demand” related T2MI were present: in those with T2MI, a slightly higher EV/Plasma ratio of 7% and EV/NEV ratio of 8% were observed. While the predominant mechanism remains passive release due to myocardial supply/demand mismatch, the higher proportion of EV-associated troponin suggests a modest contribution of active vesicular secretion, possibly triggered by ongoing ischemic stress or inflammatory activation. This aligns with the heterogeneous pathophysiology of T2MI, where myocardial injury is often driven by systemic factors such as hypoxia, anemia, or sepsis rather than direct coronary occlusion (McCarthy et al., 2021). Curiously, individuals with ESKD displayed also a relatively low EV/Plasma ratio (5%) and an EV/NEV ratio of 7%, suggesting that uremic states might favor predominantly NEV troponin release, potentially attributable to chronic subclinical myocardial injury, combined with inadequate clearance.

In the clinical stress-test cohort, plasma hs-cTnT was slightly higher at baseline (pre-exercise median ∼8.8 ng/L) than in healthy volunteers, with a lower EV contribution (EV/NEV ∼47.5%). Immediately after exercise, plasma hs-cTnT remained similar (∼8.9 ng/L), but the EV fraction rose (EV/NEV ∼73.6%), indicating a rapid, exercise-related shift toward vesicular export. This pattern likely reflects transient myocardial strain with catecholamine and shear-stress signaling rather than necrosis, and may also be influenced by the referral population (some with baseline ischemia/chronic injury). Because samples were drawn immediately post-exercise (not at the known delayed plasma troponin peak), these data emphasize mechanism (mode of release) more than the magnitude. Clinically, recognizing this EV-predominant shift after exertion may help avoid misclassification of exercise-related elevations.

Lastly, among endurance athletes and healthy controls, despite low absolute troponin levels, both groups showed EV/NEV ratios of 100%, suggesting nearly all of the circulating troponin in healthy individuals is derived from EV release. This is relevant as elevated troponin following endurance exercise is well-documented among individuals without ischemic heart disease, raising considerable anxiety at times (Conesa-Milian et al., 2023; Airaksinen, 2020; Neilan et al., 2006). The biological cause for EV release of troponin among those undergoing exercise and the clinical implications of such an observation are in need of further investigation to determine whether EV-associated troponin after exercise might support beneficial adaptive responses or simply reflect normal cardiac “housekeeping.”

Mechanistically, the meaning of EV-based troponin release from cardiomyocytes requires further understanding. In healthy individuals and endurance athletes, it may facilitate routine protein turnover by clearing misfolded or damaged proteins, similar to mitochondrial quality control mechanisms (Pan et al., 2023). In chronic disease states like CHF and HCM, persistent myocardial strain, oxidative stress, and inflammation may trigger an adaptive “toxic-waste-valve” mechanism, actively exporting troponin via EVs to prevent intracellular toxicity, a state resembling α-synuclein and tau clearance in neurodegenerative diseases (Limketkai et al., 2017). Additionally, troponin-containing EVs may function in intercellular communication, particularly in fibrotic and remodeling processes, similar to matrix metalloproteinases (MMPs) secreted via EVs to regulate tissue remodeling (Ma, 2016). Alternatively, pro-inflammatory or oxidative triggers may drive EV-mediated export, positioning EV troponin as a potential biomarker of ongoing myocardial remodeling. For example, among those with CHF (in whom a higher percentage of EV packaged troponin was observed in this analysis), change in hs-cTnT was associated with cardiac remodeling parameters (Murphy et al., 2021). Whether such changes were EV or NEV troponin-based remains to be seen but should be the focus of future analyses. If EV troponin release is an evolutionarily conserved and physiologically relevant mechanism, it may serve as a marker of chronic myocardial adaptation rather than simply myocardial injury. This could explain why EV-associated troponin levels do not correlate with NT-proBNP or plasma hs-cTnT in conditions like CHF, HCM, and ESKD. Instead of signifying acute damage, EV troponin may provide insight into ongoing myocardial remodeling, low-grade injury, or adaptive responses to chronic stress.

Clinically, the results of this study have potentially far-reaching implications as they might inform a process by which confusing troponin results might be better understood. Simultaneous quantification of EV troponin alongside traditional plasma troponin (and calculating EV/NEV ratios) might improve accuracy of the assay for acute MI: an elevated EV fraction (high EV/NEV ratio) could reinforce a picture of chronic, active stress rather than acute necrosis, whereas a low EV fraction with a sudden rise in NEV troponin (low EV/NEV ratio) might more strongly suggest an acute event. Furthermore, although this study did not specifically assess outcomes, EV troponin, and EV/NEV ratios may have prognostic value, especially because they appear to be independent of NT-proBNP and correlate poorly with standard troponin in chronic disease. This lack of correlation may make EV troponin an additional layer of risk stratification, potentially reflecting ongoing, active pathological processes such as inflammation or oxidative stress. Larger-scale studies are needed to clarify whether EV troponin trajectories can predict outcomes such as mortality, rehospitalization, or disease progression in heart failure.

## Limitations

The findings of this study should be interpreted with several limitations in mind. First, the relatively small group sizes (n = 25–31 per cohort) reduce statistical power for detecting subtle intergroup differences, particularly in subgroup analyses. Although the HCM cohort was numerically similar to others, lower available sample volumes further constrained plasma measurements. Moreover, a subset of samples from physiologic or lower-disease-burden cohorts (e.g., controls, athletes, stress-test participants) had values near the assay limit of detection, though MI and other high-troponin groups were unaffected. Second, the single-center design may limit generalizability to broader populations with differing comorbidities, demographics, or treatment practices. Correlation analyses were performed on the pooled dataset; cohort-stratified analyses were underpowered and are deferred to future, larger studies. Third, EV fractions were confirmed by Western blot and microfluidic particle sizing, supporting that troponin was encapsulated rather than free protein contamination (Supplementary Figure 2). Nonetheless, this study utilized total plasma EVs rather than cardiac-specific EV capture, which at this point remains a discovery platform not easily adaptable for large cohort studies (that have limiting amounts of plasma and larger sample sizes). Despite this, the consistent disease- and physiology-specific patterns observed strongly support biologically meaningful compartmentalization.

Western blot profiling of different troponin T/I isoforms or fragments, which may vary by disease state was not performed due to limited sample size and variability in availability of appropriate reagents. Future studies should incorporate cardiac EV–EV-specific enrichment and Western-based troponin isoform profiling (Troponin I and T ratios) to refine mechanistic insights. Fourth, the cross-sectional design limits our ability to determine whether EV-associated troponin has prognostic value or reflects dynamic changes in myocardial stress over time.

Longitudinal studies will be needed to assess whether EV troponin levels track with outcomes such as rehospitalization, mortality, or disease progression. Fifth, we did not investigate the fragmentation state of troponin within EVs. The FDA-approved hs-cTnT ELISA used in this study recognizes a stable central epitope (amino acids 125–147) that remains present in both intact and fragmented troponin molecules. Because proteolytic cleavage does not disrupt this epitope, the assay quantifies total troponin content but cannot differentiate between distinct fragment sizes or post-translationally modified forms. As proteolytic processing of troponin is increasingly understood to vary by mechanism of injury (e.g., necrosis, apoptosis, chronic stress), future work using mass spectrometry or epitope-specific immunoassays will be needed to determine whether EVs preferentially carry intact versus fragmented troponin, and whether fragmentation patterns differ across disease states. Finally, while this study highlights potential mechanistic differences in troponin secretion pathways, it does not directly assess the molecular processes governing vesicular versus free troponin release. Further research should elucidate the regulators of EV troponin trafficking and determine whether targeting EV secretion could provide new therapeutic opportunities.

## Summary

Our study reveals that EV-associated troponin is a distinct and potentially informative fraction of circulating troponin, offering new insights into its biological role and clinical significance. The lack of correlation with plasma hs-cTnT or NT-proBNP suggests that EV troponin is released through mechanisms beyond myocardial stretch or necrosis. In MI and ESRD, troponin is primarily NEV-associated, reflecting passive leakage from necrosis or impaired clearance. In AHF, myocardial strain leads to moderate EV/NEV ratios, suggesting mixed release mechanisms. In CHF and HCM, a significant proportion of troponin is packaged in EVs, indicating chronic stress-associated vesicular secretion, while in healthy controls and endurance athletes, troponin is predominantly EV-associated despite minimal plasma levels, suggesting a role in physiological turnover. These findings introduce a new perspective on troponin biology, underscoring the need for further studies to explore its compartmentalization, mechanistic pathways, prognostic value, and clinical applications in cardiovascular disease.

## Authorship Contribution

J.J., S.D., M.S., and P.G. conceived the study. J.J., S.D., M.S., and P.G. designed the research plan. A.T.R., M.W., and C.L. supervised data collection and led data interpretation. A.S., M.S., P.G., A.C.M, and R.W. optimized and performed extracellular vesicle isolation and biochemical assays. P.J. and T.O. organized and performed hs-troponin detection assays. M.S. and P.G. performed statistical analyses, prepared all figures, and wrote the main manuscript text. S.D. and J.J. provided conceptual guidance, contributed to study design, and critically revised the manuscript. J.J. additionally contributed to clinical interpretation, biomarker framework development, and critical review. All authors reviewed and approved the final manuscript.

## Data Availability

The datasets generated and analyzed during the current study contain protected health information and are therefore not publicly available. De-identified data may be accessed upon reasonable request to the corresponding author, subject to institutional approvals and compliance with Mass General Brigham IRB requirements.

## Supplementary Method: Developing and Optimizing the Troponin-Rich EV Isolation Method

To ensure efficient separation of EV-associated and non-EV (NEV) troponin, we systematically tested different isolation strategies and optimized key hydrodynamic parameters. We first compared multiple SEC column types, including qEV1 (35 nm and 70 nm), qEVoriginal (35 nm and 70 nm), and qEVsingle (35 nm and 70 nm), to determine which provided the best separation efficiency. Among these, qEV1 70 nm demonstrated the highest efficiency in isolating EV-associated troponin while minimizing free troponin contamination, making it the optimal choice for subsequent analyses.

Following column selection, we optimized the hydrodynamic parameters of isolation, including buffer volume, fraction size, fraction count, and packed column volume (PCV). The buffer volume was adjusted to ensure proper column equilibration and maximize EV elution, while the fraction size was refined to 700 µL to balance EV purity and yield. PCV was maintained at 3.5 mL to provide effective size-based separation. To confirm that the troponin detected in the EV fraction was truly vesicle-associated and not present in a free-floating form, we employed Amicon centrifugal filtration, ensuring that only encapsulated troponin was retained in the EV fraction. All fractions were tested with the bicinchoninic acid (BCA) assay for protein quantification. Because plasma contains epitopes that can appear in late SEC fractions and yield false positives, we required concordance of three independent readouts within our EV pool: (i) enrichment of canonical EV markers by Western blot (CD81, ALIX), (ii) detection of troponin by targeted proteomic analysis in those same fractions, and (iii) orthogonal confirmation of nanoparticles by Spectradyne NCS1. This convergence demonstrates that troponin resides within bona fide EVs and not as contaminating soluble protein.

All fractions were initially analyzed for EV and troponin content. Fractions 1–4 consistently contained EVs, but fraction 5 also demonstrated reproducible enrichment of EV markers and troponin, leading to their inclusion in the final EV pool. The EV fractions were pooled and validated for EV markers (CD81, ALIX) in different conditions using Western blot, as observed in **Supplementary Figure 2.** To characterize vesicle elution on the Izon qEV35 format used during method development, we processed 10 independently pooled plasma samples and collected 18 fractions per run; each sample was measured in triplicate on the Spectradyne NCS1. Nanovesicle concentrations (particles/mL) peaked in fractions 1–4 (≈3.5–4.2 × 10¹¹ particles/mL) and declined sharply after fraction 5, with a lower level signal in fractions 6–18 (10 –10¹ particles/mL). As mentioned earlier, these data met the criteria: the coexistence of highly enriched canonical EV markers, Troponin, and high counts of nanoparticles, and supported the inclusion of fractions 1–5 in the EV pool and the designation of fractions 6–20 as NEV.

The NEV fraction was collected from fractions 6–20, as these primarily contained free protein, non-vesicular components, and significantly lower counts of measured nanoparticles. This refined protocol ensures robust and reproducible isolation of EV-associated troponin, enabling accurate downstream analysis of its biological relevance.

## SUPPLEMENTARY FIGURES

**Supplementary Figure 1.**
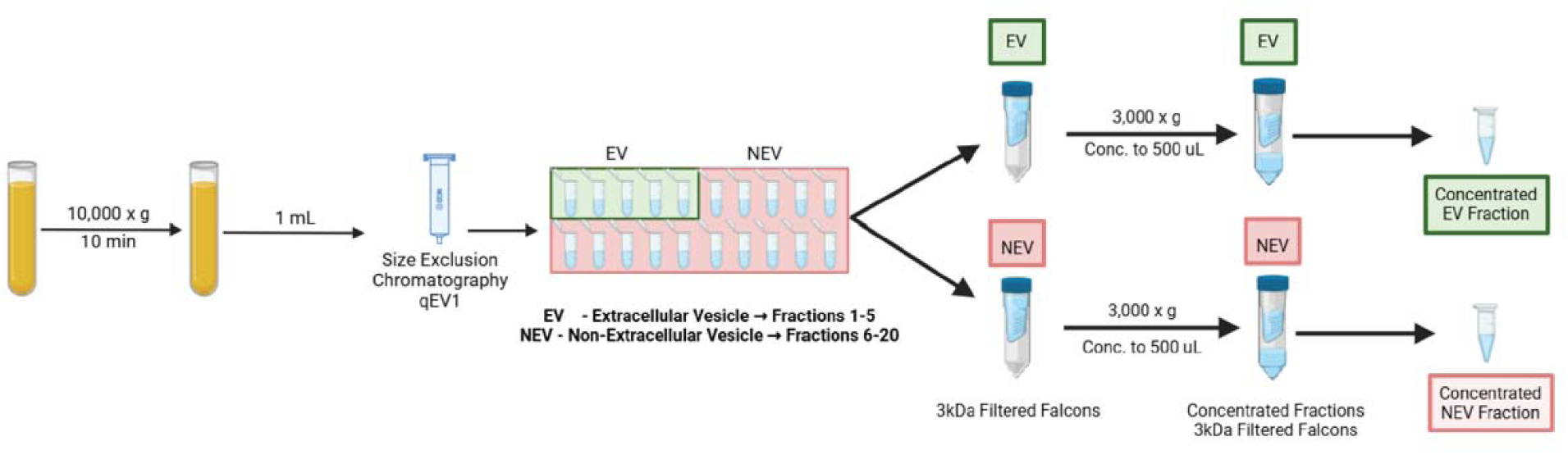
Schematic representation of the methodology used to obtain EV (Extracellular Vesicle) Fractions and NEV (Non-Extracellular Vesicle) Fractions from 1mL of plasma using size exclusion chromatography.

**Supplementary Figure 2.**
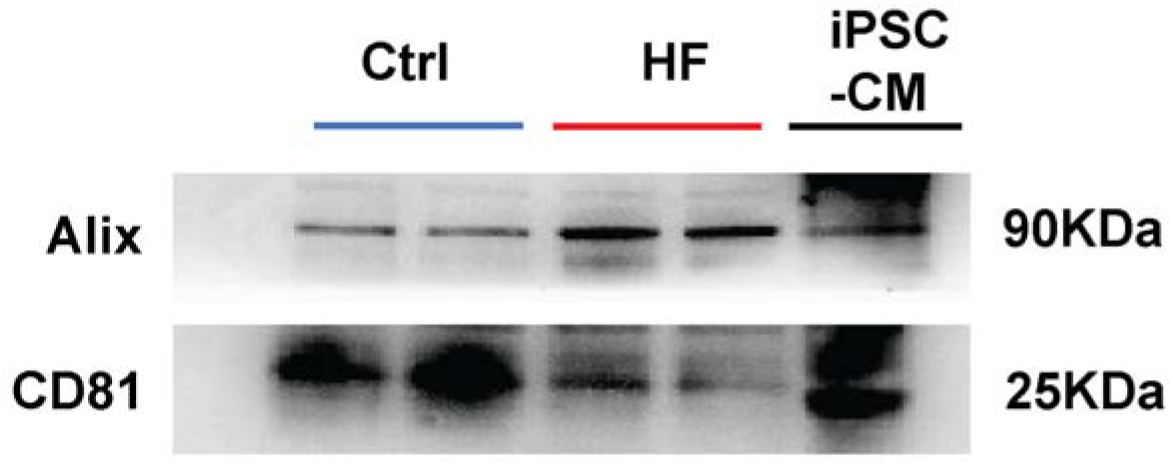
Western Blot analysis showing the EV markers **Alix** (an EV-cargo protein) and **CD81** (a tetraspanin, EV-membrane marker) on plasma EVs isolated from Control (Ctrl), Heart Failure (HF) and induced Pluripotent Stem Cell derived cardiomyocytes (iPSC-CMs).

**Supplementary Figure 3.**
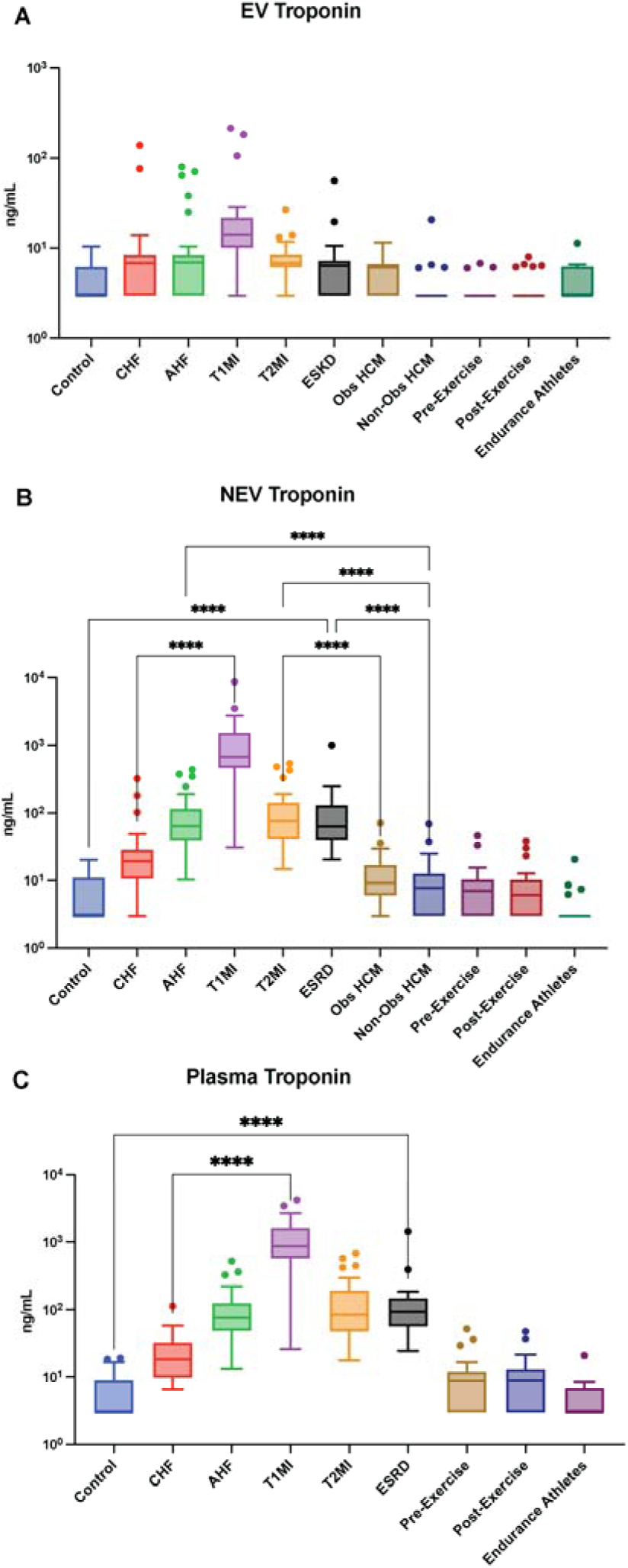
**A.** Plasma Troponin **B.** Plasma Troponin **C.** Plasma Troponin measurements in different disease groups, including Chronic Heart Failure (CHF), Acute Heart Failure (AHF), Type 1 Myocardial Infarction (T1MI), Type 2 Myocardial Infarction (T2MI), End- Stage Renal Disease (ESRD), Obstructive Hypertrophic Cardiomyopathy (Obs-HCM), Non- Obstructive Hypertrophic Cardiomyopathy (Non Obs-HCM), along Healthy Controls, Endurance Athletes, Pre and Post-Exercise. The most significant comparisons are shown with * p<0.0332 ** p<0.0021 ***p<0.0002****p<0.00001where p is calculated using Kruskal-Wallis test using GraphPad Prism.

**Supplementary Figure 4.**
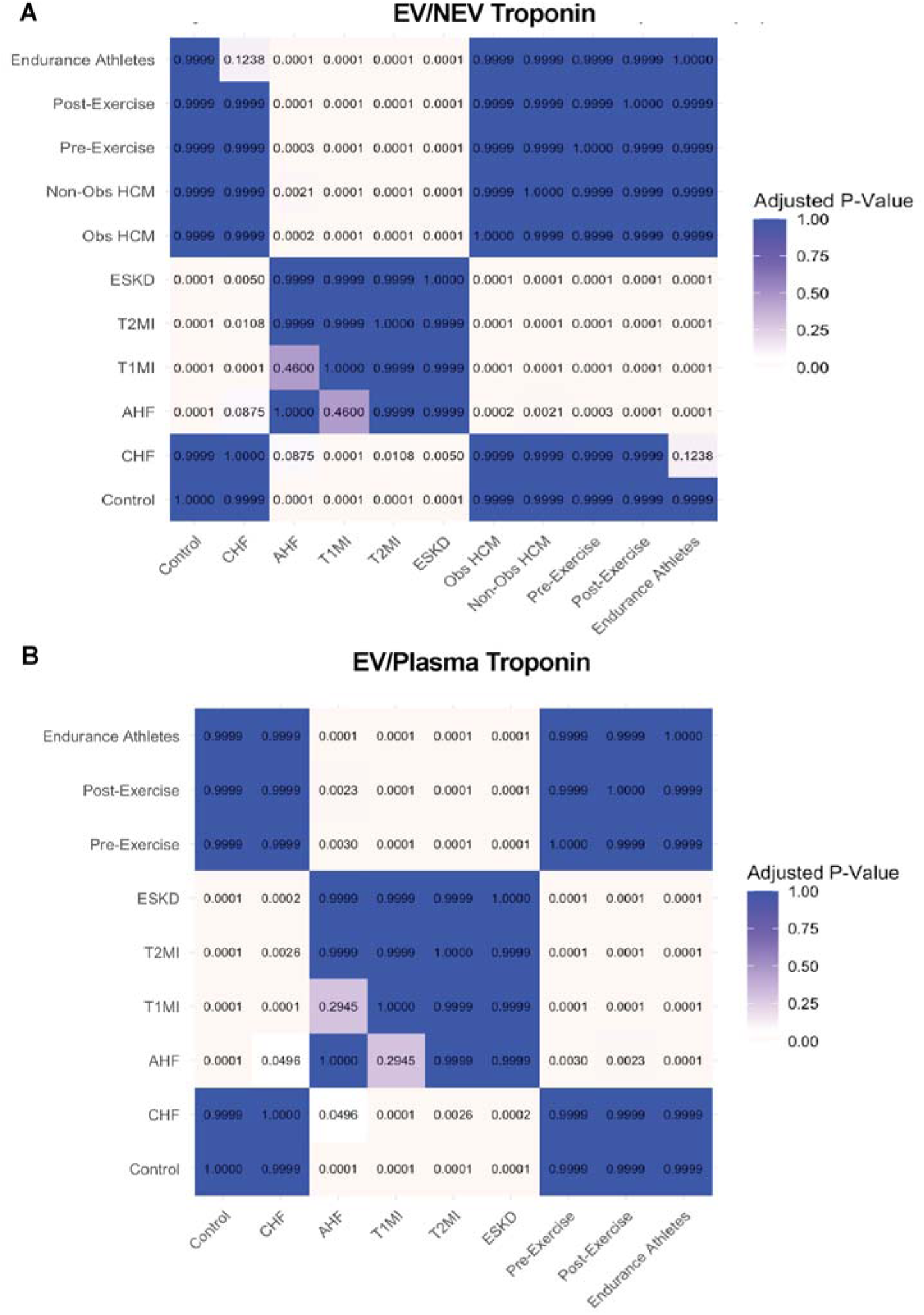
Adjusted p-value matrix (Dunn’s test) for EV/NEV Troponin (**A**) and EV/Plasma Troponin (**B**) comparison between the different pathological groups, including Endurance Athletes, Healthy Controls, Pre- and Post-Exercise, Hypertrophic Cardiomyopathy (HCM), Chronic Heart Failure (CHF), Acute Heart Failure (AHF), Type 1 Myocardial Infarction (T1MI), Type 2 Myocardial Infarction (T2MI), and End-Stage Renal Disease (ESRD). EV indicated by Extracellular Vesicle Fraction, and NEV indicated by Non-Extracellular Vesicle Fraction of 1mL Plasma isolated using size exclusion chromatography.

## SUPPLEMENTARY TABLE

**Supplementary Table:**
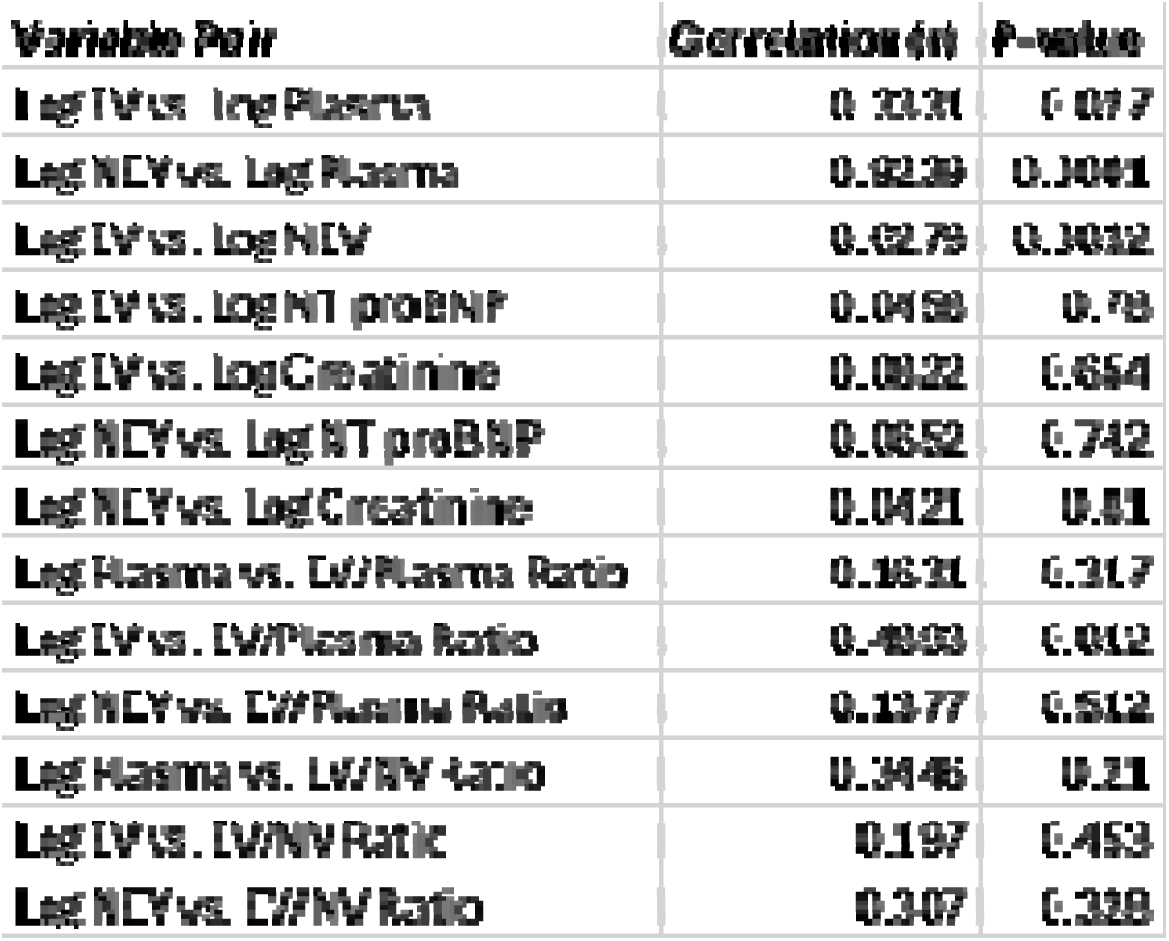
Pearson correlation coefficients (r) and exact p-values for pairwise comparisons of log-transformed variables, including EV, NEV, and Plasma troponin levels, as well as NT-proBNP, creatinine, and derived ratios (EV/Plasma and EV/NV).

| **Variable Pair** | **Correlation (r)** | **P-value** |
| --- | --- | --- |
| **Log IV vs. Log Plasma** | **0.3331** | **0.017** |
| **Log NEV vs. Log Plasma** | **0.0239** | **0.3041** |
| **Log IV vs. Log NEV** | **0.0279** | **0.3012** |
| **Log IV vs. Log NT proBNP** | **0.0458** | **0.78** |
| **Log IV vs. Log Creatinine** | **0.0022** | **0.654** |
| **Log NEV vs. Log NT proBNP** | **0.0652** | **0.742** |
| **Log NEV vs. Log Creatinine** | **0.0421** | **0.81** |
| **Log Plasma vs. EV/Plasma Ratio** | **0.1631** | **0.317** |
| **Log IV vs. EV/Plasma Ratio** | **0.4003** | **0.012** |
| **Log NEV vs. EV/Plasma Ratio** | **0.1377** | **0.512** |
| **Log Plasma vs. EV/NV Ratio** | **0.3446** | **0.21** |
| **Log IV vs. EV/NV Ratio** | **0.197** | **0.453** |
| **Log NEV vs. EV/NV Ratio** | **0.307** | **0.328** |

## References

Airaksinen, K. E. J. (2020). Cardiac Troponin Release After Endurance Exercise: Still Much to Learn. Journal of the American Heart Association, 9(4), e015912. 10.1161/JAHA.120.015912

Akbas, T. (2024). Elevated Cardiac Troponin Levels as a Predictor of Increased Mortality Risk in Non-Cardiac Critically Ill Patients Admitted to a Medical Intensive Care Unit. Journal of Clinical Medicine, 13(20), Article 20. 10.3390/jcm13206025

Conesa-Milian, E., Cirer-Sastre, R., Hernández-González, V., Legaz-Arrese, A., Corbi, F., & Reverter-Masia, J. (2023). Cardiac Troponin Release after Exercise in Healthy Young Athletes: A Systematic Review. *Healthcare (Basel*, Switzerland), 11(16), 2342. 10.3390/healthcare11162342

Eggers, K. M., Jernberg, T., & Lindahl, B. (2019). Cardiac Troponin Elevation in Patients Without a Specific Diagnosis. Journal of the American College of Cardiology, 73(1), 1–9. 10.1016/j.jacc.2018.09.082

Ilva, T. J., Eskola, M. J., Nikus, K. C., Voipio-Pulkki, L.-M., Lund, J., Pulkki, K., Mustonen, H., Niemelä, K. O., Karhunen, P. J., & Porela, P. (2010). The etiology and prognostic significance of cardiac troponin I elevation in unselected emergency department patients. The Journal of Emergency Medicine, 38(1), 1–5. 10.1016/j.jemermed.2007.09.060

Januzzi, J. L., Bamberg, F., Lee, H., Truong, Q. A., Nichols, J. H., Karakas, M., Mohammed, A. A., Schlett, C. L., Nagurney, J. T., Hoffmann, U., & Koenig, W. (2010). High-sensitivity troponin T concentrations in acute chest pain patients evaluated with cardiac computed tomography. Circulation, 121(10), 1227–1234. 10.1161/CIRCULATIONAHA.109.893826

Januzzi, J. L., Filippatos, G., Nieminen, M., & Gheorghiade, M. (2012). Troponin elevation in patients with heart failure: On behalf of the third Universal Definition of Myocardial Infarction Global Task Force: Heart Failure Section. European Heart Journal, 33(18), 2265–2271. 10.1093/eurheartj/ehs191

Jeremias, A., & Gibson, C. M. (2005). Narrative review: Alternative causes for elevated cardiac troponin levels when acute coronary syndromes are excluded. Annals of Internal Medicine, 142(9), 786–791. 10.7326/0003-4819-142-9-200505030-00015

Kakihana, Y., Ito, T., Nakahara, M., Yamaguchi, K., & Yasuda, T. (2016). Sepsis-induced myocardial dysfunction: Pathophysiology and management. Journal of Intensive Care, 4(1), 22. 10.1186/s40560-016-0148-1

Lennon, K. M., Saftics, A., Abuelreich, S., Sahu, P., Lehmann, H. I., Maddox, A. L., Bagabas, R., Januzzi, J. L., Van Keuren-Jensen, K., Shah, R., Das, S., & Jovanovic-Talisman, T. (2022). Cardiac troponin T in extracellular vesicles as a novel biomarker in human cardiovascular disease. Clinical and Translational Medicine, 12(8), e979. 10.1002/ctm2.979

Limketkai, B. N., Parian, A. M., Chen, P.-H., & Colombel, J.-F. (2017). Treatment With Biologic Agents Has Not Reduced Surgeries Among Patients With Crohn’s Disease With Short Bowel Syndrome. Clinical Gastroenterology and Hepatology: The Official Clinical Practice Journal of the American Gastroenterological Association, 15(12), 1908–1914.e2. 10.1016/j.cgh.2017.06.040

Ma, X. J. (2016). [Guidelines for Nontuberculous mycobacterial and its’ clinical applicability]. Zhonghua Nei Ke Za Zhi, 55(4), 264–266. 10.3760/cma.j.issn.0578-1426.2016.04.003

McCarthy, C. P., Kolte, D., Kennedy, K. F., Vaduganathan, M., Wasfy, J. H., & Januzzi, J. L. (2021). Patient Characteristics and Clinical Outcomes of Type 1 Versus Type 2 Myocardial Infarction. Journal of the American College of Cardiology, 77(7), 848–857. 10.1016/j.jacc.2020.12.034

McCarthy, C. P., Wasfy, J. H., & Januzzi, J. L. (2024). Is Myocardial Infarction Overdiagnosed? JAMA, 331(19), 1623–1624. 10.1001/jama.2024.5235

Mohammed, A. A., & Januzzi, J. L. (2010). Clinical applications of highly sensitive troponin assays. Cardiology in Review, 18(1), 12–19. 10.1097/CRD.0b013e3181c42f96

Murphy, S. P., Prescott, M. F., Maisel, A. S., Butler, J., Piña, I. L., Felker, G. M., Ward, J. H., Williamson, K. M., Camacho, A., Kandanelly, R. R., Solomon, S. D., & Januzzi, J. L. (2021). Association Between Angiotensin Receptor-Neprilysin Inhibition, Cardiovascular Biomarkers, and Cardiac Remodeling in Heart Failure With Reduced Ejection Fraction. Circulation. Heart Failure, 14(6), e008410. 10.1161/CIRCHEARTFAILURE.120.008410

Neilan, T. G., Januzzi, J. L., Lee-Lewandrowski, E., Ton-Nu, T.-T., Yoerger, D. M., Jassal, D. S., Lewandrowski, K. B., Siegel, A. J., Marshall, J. E., Douglas, P. S., Lawlor, D., Picard, M. H., & Wood, M. J. (2006). Myocardial injury and ventricular dysfunction related to training levels among nonelite participants in the Boston marathon. Circulation, 114(22), 2325–2333. 10.1161/CIRCULATIONAHA.106.647461

Pan, K.-H., Chang, H., & Yang, W. Y. (2023). Extracellular release in the quality control of the mammalian mitochondria. Journal of Biomedical Science, 30(1), 85. 10.1186/s12929-023-00979-3

Park, K. C., Gaze, D. C., Collinson, P. O., & Marber, M. S. (2017). Cardiac troponins: From myocardial infarction to chronic disease. Cardiovascular Research, 113(14), 1708–1718. 10.1093/cvr/cvx183

Tanindi, A., & Cemri, M. (2011). Troponin elevation in conditions other than acute coronary syndromes. Vascular Health and Risk Management, 7, 597–603. 10.2147/VHRM.S24509

Thygesen, K., Alpert, J. S., Jaffe, A. S., Chaitman, B. R., Bax, J. J., Morrow, D. A., White, H. D., & Executive Group on behalf of the Joint European Society of Cardiology (ESC)/American College of Cardiology (ACC)/American Heart Association (AHA)/World Heart Federation (WHF) Task Force for the Universal Definition of Myocardial Infarction. (2018). Fourth Universal Definition of Myocardial Infarction (2018). Journal of the American College of Cardiology, 72(18), 2231–2264. 10.1016/j.jacc.2018.08.1038

